# Resolving Diagnostic Discordance in Group 2 Pulmonary Hypertension Through Staged Physiologic Testing: Insights From PVDOMICS

**DOI:** 10.64898/2026.06.04.26354961

**Authors:** Franz P. Rischard, Mateo Mendoza, Michael Insel, Gerald J. Beck, Serpil C. Erzurum, Robert P. Frantz, J. Emanuel Finet, Paul M. Hassoun, Anna R. Hemnes, Nicholas S. Hill, Evelyn M. Horn, Jane A. Leopold, Stephen C. Mathai, Reena Mehra, Yogesh N.V. Reddy, Erika B. Rosenzweig, David M. Systrom, W H Wilson Tang, Aaron B. Waxman, Barry A. Borlaug, the PVDOMICS Study Group

## Abstract

**Background:** World Symposium on Pulmonary Hypertension (WSPH) Group 2 pulmonary hypertension (PH) is a clinically integrated phenotype attributed to left heart disease, whereas pre- versus post-capillary classification is operationalized primarily by pulmonary capillary wedge pressure (PCWP). Although current recommendations emphasize contextual interpretation and provocative testing for intermediate PCWP values, the relationship between PCWP-based classification and underlying phenotype has not been systematically evaluated. We aim to quantify phenotype–hemodynamic discordance across the PCWP spectrum and evaluate a staged physiology-guided framework incorporating inhaled nitric oxide (iNO), ventricular geometry, and provocative testing.

**Methods:** We studied 1,032 participants from the NHLBI-sponsored PVDOMICS cohort with multidisciplinary adjudicated phenotypes integrating clinical, imaging, physiologic, and hemodynamic data. Stage-specific PCWP thresholds classified pre- versus post-capillary physiology at rest, during iNO, and during provocation (fluid challenge or invasive cardiopulmonary exercise testing [iCPET]). Echocardiographic right ventricular-to-left ventricular (RV/LV) ratio was evaluated as a marker of ventricular interdependence. Restricted cubic spline and staged concordance analyses defined certainty-based PCWP ranges and incremental diagnostic yield.

**Results:** Adjudicated Group 2 PH was present in 37.0% of participants. Resting PCWP demonstrated good discrimination (AUC 0.86), but substantial bidirectional phenotype–hemodynamic discordance persisted across intermediate PCWP ranges. At a resting PCWP of 12 mmHg, 25% of participants classified as pre-capillary had adjudicated Group 2 PH, whereas at 18 mmHg, 35% classified as post-capillary remained discordant non–Group 2. Concordance did not approach 90% until PCWP values were <9 mmHg or >24 mmHg. Dynamic testing incrementally improved concordance within these overlap zones. Nearly half of adjudicated Group 2 PH participants (46.5%) were not identified by resting PCWP alone; incorporation of iNO and provocative testing increased cumulative Group 2 identification by 63.4% and improved sensitivity from 79.9% to 83.7%. Model discrimination improved from an AUC of 0.863 to 0.908 (likelihood-ratio P<0.001). iNO increased PCWP in discordant Pre/G2 participants, unmasking latent left-sided limitation, while lowering PCWP in discordant Post/NonG2 participants, consistent with ventricular interdependence. RV/LV ratio ≥0.94 reduced discordant Post/NonG2 classification by 70.5%, and incorporation of PCWP/cardiac output slope improved physiologic specificity during exercise.

**Conclusions:** Group 2 PH is a dynamic, load-dependent phenotype inadequately characterized by resting PCWP alone. Intermediate PCWP values represent continuous probabilities of bidirectional discordance rather than discrete diagnostic states. A staged physiology-guided approach integrating iNO, ventricular geometry, and provocative testing improves concordance between hemodynamic classification and clinically integrated phenotype assignment.

**Clinical Perspective:** *What Is New?:*

- In the deeply phenotyped PVDOMICS cohort, resting pulmonary capillary wedge pressure demonstrated good overall discrimination for adjudicated Group 2 (left heart disease) pulmonary hypertension, yet intermediate values frequently either concealed latent left-heart disease or overclassified patients without intrinsic left-heart disease
- Resting pulmonary capillary wedge pressure should be viewed as a continuous probability signal rather than a binary threshold, allowing additional physiologic testing to be targeted according to the degree of diagnostic uncertainty a clinician is willing to accept.
- A staged physiology-guided approach incorporating inhaled nitric oxide, ventricular geometry, and provocative testing improved concordance with adjudicated PH category and pre or post-capillary classification.

*What Are the Clinical Implications?:*

- Pre- versus post-capillary classification should be interpreted within the broader clinical and physiologic context rather than relying on a single resting pulmonary capillary wedge pressure threshold.
- Intermediate pulmonary capillary wedge pressure values should prompt consideration of additional physiologic evaluation, with inhaled nitric oxide providing a practical intermediate step and provocative testing providing the greatest incremental diagnostic yield.
- Exercise pulmonary capillary wedge pressure/cardiac output slope and markers of ventricular interdependence may provide complementary information for resolving uncertainty when resting and dynamic hemodynamics are discordant.

## Introduction

Pulmonary hypertension (PH) classification within the World Symposium on Pulmonary Hypertension (WSPH) framework is inherently multiparametric, integrating clinical history, comorbidities, imaging, pulmonary function testing, and invasive hemodynamics to define disease phenotype and guide management.^1,2^ Within this framework, WSPH Group 2 PH refers to PH associated with left heart disease (LHD), the most common cause of PH and a clinically important distinction because pulmonary vasoactive therapies may be ineffective or potentially harmful.^3–6^ Because direct phenotypic assignment of LHD can be challenging in clinical practice, invasive hemodynamic classification is frequently used to operationalize the distinction between PH associated with left-heart pathology versus intrinsic pulmonary vascular disease.

This hemodynamic framework classifies PH as pre- or post-capillary largely according to pulmonary capillary wedge pressure (PCWP), with post-capillary PH defined by PCWP >15 mmHg.^1,7^ However, WSPH statements emphasize that PCWP should be interpreted within clinical context because waveform methodology, loading conditions, ventricular interaction, and occult left-heart dysfunction may influence classification.^8–10^ Current recommendations suggest that patients with intermediate PCWP values (e.g. 12–18 mmHg)^1,8^ may require integrated clinical interpretation and provocative testing, yet these recommendations have not been systematically evaluated across the spectrum of pulmonary vascular disease. As a result, uncertainty persists regarding when PCWP-based hemodynamic classification accurately reflects underlying Group 2 PH phenotype and when additional physiologic testing is required to resolve phenotype–hemodynamic discordance.

Discordance between resting PCWP-based classification and clinically integrated phenotype assignment may arise in two physiologically distinct directions. Some patients with Group 2 PH demonstrate normal or borderline resting PCWP despite impaired left atrial or ventricular reserve, resulting in latent post-capillary physiology that becomes apparent only during dynamic testing.^11^ Conversely, other patients exhibit elevated resting PCWP despite absence of intrinsic left-heart disease, potentially reflecting ventricular interdependence, pericardial constraint, or right ventricular pressure overload rather than true post-capillary PH.^8,9^ The relative contribution of these bidirectional discordance profiles across the spectrum of disease, and the extent to which dynamic testing improves phenotype–hemodynamic concordance, remain incompletely defined.

The NHLBI-sponsored The Redefining Pulmonary Hypertension through Pulmonary Vascular Disease Phenomics (PVDOMICS) cohort provides a unique opportunity to address this gap through deep phenotyping, multidisciplinary adjudication, and protocolized staged hemodynamic assessment including rest, inhaled nitric oxide (iNO), and provocative testing.^12,13^ We therefore evaluated concordance and discordance between PCWP-based hemodynamic classification and adjudicated Group 2 PH assignment across staged physiologic testing to define where hemodynamic classification succeeds, where it fails, and how dynamic testing may refine physiologic interpretation.

## Methods

### Study Design and Population

The PVDOMICS study (NCT02980887), funded by the National Heart, Lung, and Blood Institute and the Pulmonary Hypertension Association, enrolled participants across seven US centers between November 30, 2016, and October 18, 2019. PVDOMICS was designed to characterize pulmonary vascular disease using standardized invasive hemodynamics, imaging, and multimodal clinical phenotyping across World Symposium on Pulmonary Hypertension (WSPH) Groups 1–5, including participants with and without resting PH.

Initial WSPH classification was performed by site principal investigators using integrated clinical assessment. Subjects with more than one WSPH Group (mixed phenotypes) underwent multidisciplinary adjudication with cardiology and pulmonology representation using clinical history, imaging, hemodynamics, and comorbidity profile. Group 2 PH included participants with primary, secondary, or tertiary Group 2 designation. WSPH Group 5 and healthy controls were excluded. Detailed adjudication procedures are provided in the Supplemental Methods.

Stage-specific analyses used paired datasets between sequential testing stages (rest→inhaled nitric oxide [iNO]; iNO→provocation). Longitudinal hemodynamic analyses incorporated all available measurements from participants contributing ≥1 testing stage.

### Invasive Hemodynamic Testing

All invasive procedures were performed according to standardized PVDOMICS protocols (Supplemental Methods). Resting right heart catheterization was performed in the supine position with measurement of right atrial pressure (RAP), mean pulmonary artery pressure (mPAP), pulmonary arterial wedge pressure (PCWP), and cardiac output by thermodilution. Respiratory-averaged PCWP was used to preserve physiologic continuity across dynamic testing stages, including exercise, where respiratory averaging is preferred.^14^ A supplementary sensitivity analysis using end-expiratory PCWP measurements^15^ was also performed for resting, iNO, and fluid challenge stages. Derived indices included pulmonary vascular resistance (PVR) and transmural pressure (TMP), calculated as PCWP−RAP.^16^

Following baseline measurements, iNO was administered at 40 ppm for 10 minutes with repeat hemodynamics obtained after stabilization. Provocative testing consisted of either fluid challenge or invasive cardiopulmonary exercise testing (iCPET), according to protocol availability and participant suitability. Fluid challenge consisted of rapid isotonic saline infusion (500 mL over 10 minutes) with repeat post-infusion hemodynamics. iCPET was performed using cycle ergometry with simultaneous invasive hemodynamic and breath-by-breath gas exchange assessment. Detailed provocative testing protocols are provided in the Supplemental Methods.

### Hemodynamic Classification Across Physiologic States

Pre- versus post-capillary hemodynamic profile was defined separately at each testing stage for PH subjects using stage-specific PCWP thresholds reflecting physiologic context rather than a single static cutoff. Post-capillary physiology was defined as PCWP >15 mmHg at rest or during iNO,^1,7^ >18 mmHg during fluid challenge,^17,18^ and ≥20 mmHg (upright, 2 centers)^19^ or ≥25 mmHg (supine, 1 center)^17,20^ during iCPET, consistent with established physiologic and guideline-supported thresholds. To improve physiologic specificity during exercise, a modified iCPET definition additionally requiring a PCWP–cardiac output slope >2 mmHg·L⁻¹·min⁻¹ was evaluated using the methods of Eisman et al.^18,21,22^

Because ventricular interaction may contribute to discordant post-capillary hemodynamic profiles through load-dependent wedge elevation, echocardiographic right ventricular (RV) and left ventricular (LV) end-diastolic chamber dimensions were measured to derive an RV/LV ratio as a structural surrogate of ventricular interdependence. Detailed echocardiographic methods are provided in the Supplemental Methods.

### Concordance Between Adjudicated Clinical Phenotype and Hemodynamic Classification

All subjects underwent standardized clinical adjudication for the presence or absence of left heart disease (LHD), independent of pulmonary hypertension (PH) status. This adjudication framework generated four mutually exclusive clinical phenotype groups: (1) LHD without PH, (2) Group 2 pulmonary hypertension (G2PH), (3) non-LHD without PH, and (4) non-G2PH (**Figure 1, Panel A**).

**Figure 1.**
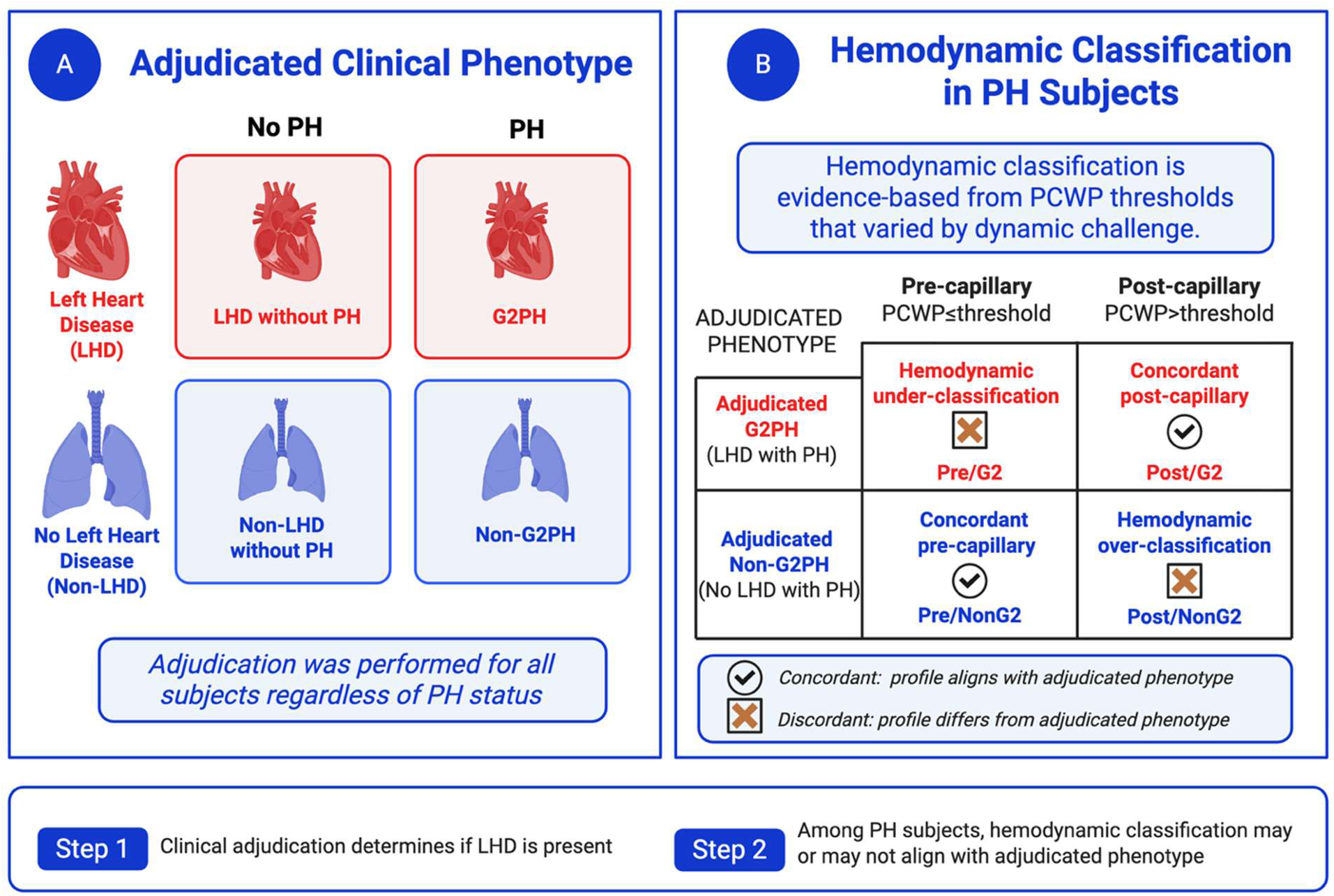
Conceptual framework for phenotype–hemodynamic concordance. (**A**) Participants are classified along two independent axes: adjudicated phenotype (Group 2 [left heart disease] vs non–Group 2) and pulmonary hypertension (PH) status. Group 2 PH includes participants both with (G2PH) and without PH (G2/no PH). (**B**) Resting hemodynamic classification (pre- vs post-capillary, based on PCWP) is evaluated against adjudicated phenotype, independent of PH status. Cross-classification defines concordant categories—pre-capillary/non–Group 2 and post-capillary/Group 2 and discordant categories—pre-capillary/Group 2 and post-capillary/non–Group 2.

Because pre- versus post-capillary terminology represents a PH-specific hemodynamic construct, concordance analyses were restricted to subjects with PH. Accordingly, hemodynamic classification was not applied to subjects without PH (**Figure 1, Panel B**).

Among PH subjects, hemodynamic classification was compared against the adjudicated clinical phenotype framework. Subjects with clinically adjudicated LHD and PH were classified as adjudicated G2PH, whereas PH subjects without clinically adjudicated LHD were classified as adjudicated non-G2PH. Hemodynamic classification was assigned using evidence-based pulmonary capillary wedge pressure (PCWP) thresholds defined according to physiologic condition or dynamic challenge, as described above.

Concordance categories within PH subjects were defined as follows:

- **Concordant post-capillary physiology:** post-capillary hemodynamics in subjects with adjudicated G2PH (Post/G2)
- **Discordant hemodynamic over-classification:** post-capillary hemodynamics in subjects with adjudicated non-G2PH (Post/NonG2)
- **Discordant hemodynamic under-classification:** pre-capillary hemodynamics in subjects with adjudicated G2PH (Pre/G2)
- **Concordant pre-capillary physiology:** pre-capillary hemodynamics in subjects with adjudicated non-G2PH (Pre/NonG2)

This framework evaluated the degree to which hemodynamic profiles aligned with the adjudicated clinical phenotype hierarchy rather than treating resting hemodynamics as an independent diagnostic gold standard. Stage-wise shifts in concordance were evaluated following iNO and provocative testing.

### Statistical Analysis

Continuous variables are presented as median [interquartile range] and compared using Wilcoxon rank-sum or Kruskal-Wallis testing, as appropriate. Categorical variables are presented as counts (%) and compared using Pearson chi-square or Fisher exact testing. Repeated-measures hemodynamics across physiologic stages were analyzed using generalized linear mixed-effects models with subject-level random intercepts, including testing stage, Group 2 PH, and their interaction. Logistic regression models were used to evaluate stage-wise operating characteristics relative to adjudicated phenotype assignment, with nested model comparisons performed using likelihood ratio testing and area under the receiver operating characteristic curve (AUC) comparisons using DeLong testing.^23^ For staged analyses, exercise and fluid challenge were grouped as a unified provocation stage to evaluate the incremental contribution of dynamic reserve physiology beyond resting and iNO assessment. This approach was based on the shared physiologic objective of increasing LV filling requirements and on directionally similar effects on phenotype–hemodynamic concordance observed across modalities. To assess potential modality-specific differences, exercise and fluid challenge cohorts were additionally analyzed separately, including stage-wise hemodynamics, concordance distributions, and nested discrimination analyses. Dual-threshold overlap analyses were performed to identify regions of high phenotype–hemodynamic concordance and intermediate overlap. Additional spline analyses, and classification and regression tree analyses are provided in the Supplemental Methods. All analyses were performed using R version 4.3 and SPSS version 31. Two-sided p-values <0.05 were considered statistically significant.

## Results

### Study Population and Analytic Cohort

Among 1,195 participants enrolled in PVDOMICS, 1,058 met eligibility criteria for the present analysis after exclusion of healthy controls, WSPH Group 5 participants, and indeterminate phenotypes (**Supplemental Figure S1**). Resting hemodynamic classification was available in 1,032 participants, paired rest→iNO data in 767 participants, and paired iNO→provocation data in 635 participants (fluid challenge n=375; iCPET n=260).

The cohort preserved the broad physiologic spectrum characteristic of PVDOMICS, including participants with and without resting pulmonary hypertension across adjudicated phenotypes. Isolated LHD was uncommon; mixed phenotypes, most commonly overlapped with lung disease (**Supplemental Table S1**). **Supplemental Figure 2** summarizes cohort stratification according to the presence or absence of resting pulmonary hypertension (PH) and adjudicated left heart disease (LHD) phenotype. Eight-hundred thirty-one subjects (80.5%) met criteria for resting PH (mPAP >20 mmHg), whereas 201 (19.5%) did not have resting PH (mPAP ≤20 mmHg). Among participants without resting PH, LHD phenotype was present in 65 (31.4%). These subjects had higher prevalence of HFrEF, valvular heart disease, and HFpEF than subjects without LHD despite similar resting hemodynamics (**Supplemental Table S2**).

As expected, adjudicated Group 2 PH participants were older and demonstrated greater cardiometabolic comorbidity burden, including higher prevalence of hypertension, diabetes, HFpEF, HFrEF, and valvular heart disease (**Table 1**). Group 2 participants also exhibited higher H2FPEF scores, larger left atrial volumes, greater LV mass, and higher resting and dynamic PCWP values. In contrast, non–Group 2 PH participants demonstrated higher transpulmonary gradients, diastolic pressure gradients, pulmonary vascular resistance, and greater right-sided chamber predominance. Across physiologic stages, Group 2 PH participants demonstrated progressive increases in PCWP and RAP with iNO and provocative testing, whereas non–Group 2 participants retained higher pulmonary vascular loading and lower wedge pressures despite dynamic perturbation (**Figure 2**).

**Figure 2.**
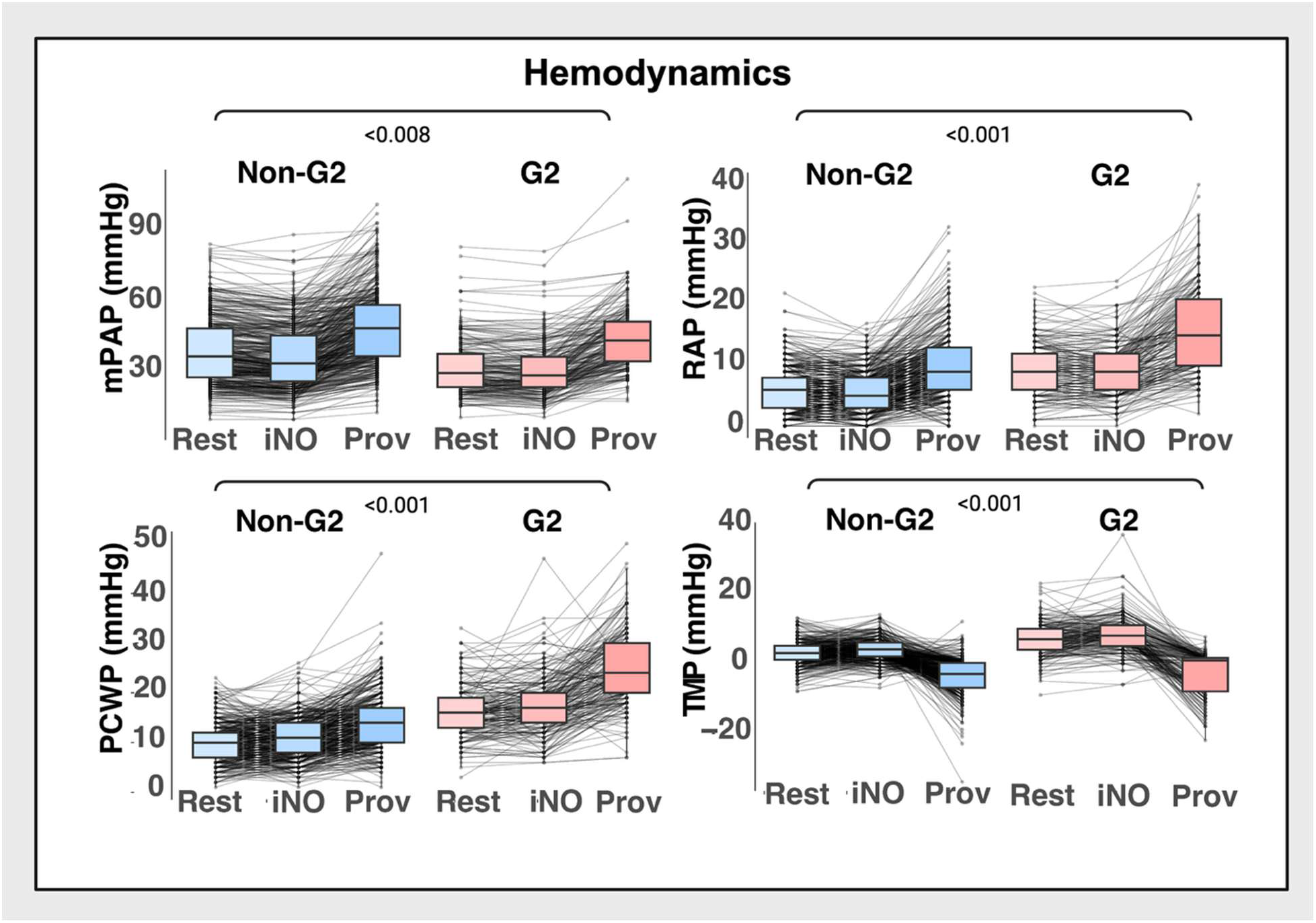
Stage-dependent hemodynamic responses to inhaled nitric oxide and provocation stratified by Group 2 PH status. **Top panels:** Mean pulmonary artery pressure (mPAP, left) and right atrial pressure (RAP, right) across rest, inhaled nitric oxide (iNO), and provocation are shown for participants without Group 2 PH (Non-G2) and those with adjudicated Group 2 PH (G2). **Bottom panels:** Pulmonary capillary wedge pressure (PCWP, left) and transmural pressure (TMP, right) across testing stages. In G2 disease, PCWP increased stepwise from rest to iNO and further with provocation, whereas Non-G2 participants demonstrated smaller absolute shifts. Transmural pressure declined with provocation in both groups but remained higher in G2 participants. **Across panels:** Thin gray lines represent within-subject trajectories across testing stages; boxplots display median and interquartile range. *P* values reflect Group × Stage interaction terms derived from generalized linear mixed-effects models, testing whether hemodynamic responses across rest, iNO, and provocation differed by Group 2 disease status. **Abbreviations:** G2, Group 2 PH; iNO, inhaled nitric oxide; mPAP, mean pulmonary artery pressure; PCWP, pulmonary capillary wedge pressure; RAP, right atrial pressure; TMP, transmural pressure; Prov, provocation (fluid challenge or invasive cardiopulmonary exercise testing).

**Table 1.**
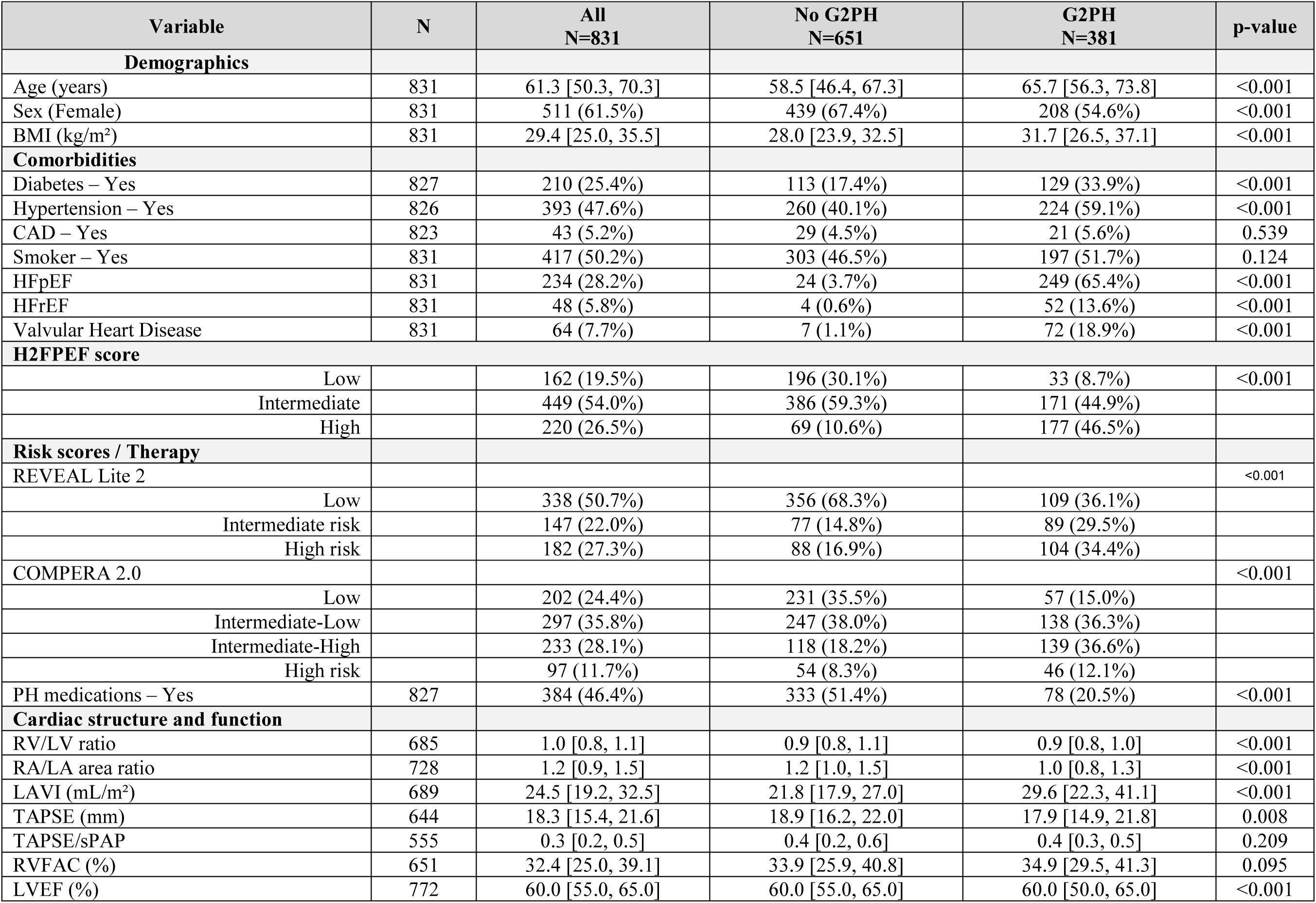
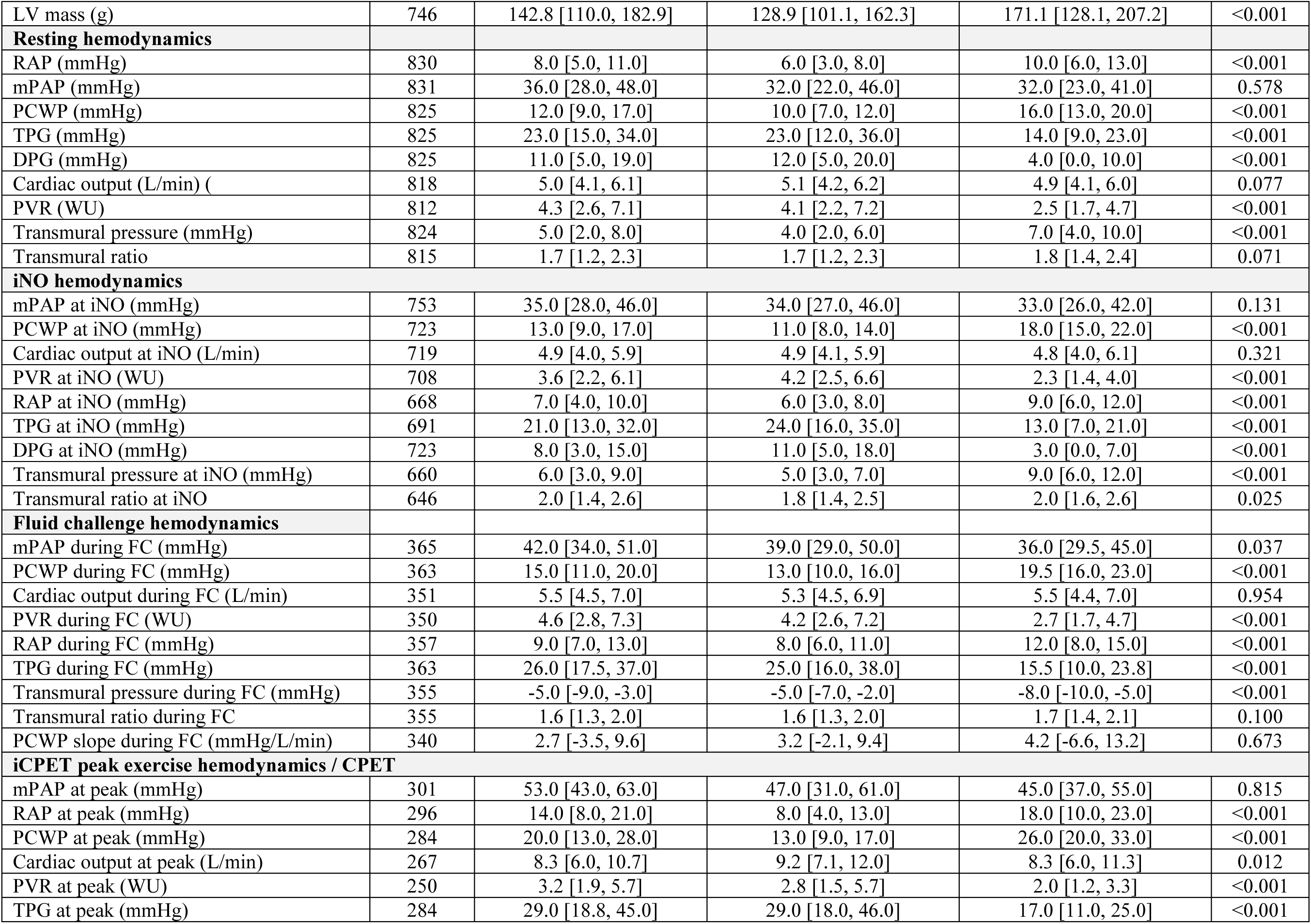
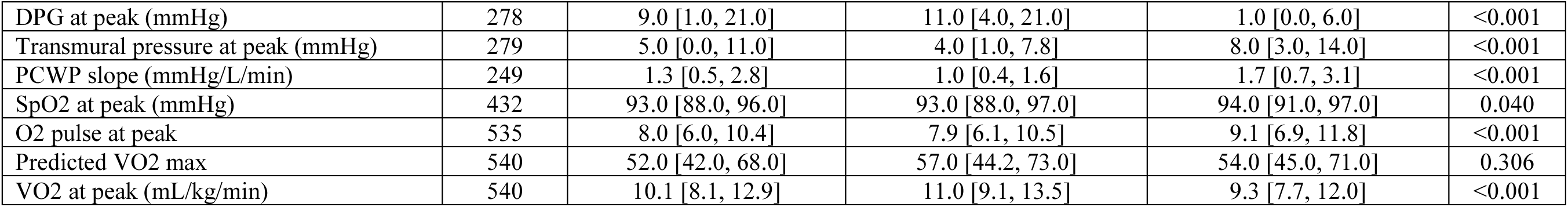
Baseline clinical characteristics, cardiac structure and function, and invasive hemodynamics stratified by adjudicated G2PH status. Demographics, comorbidities, risk scores, cardiac imaging metrics, and invasive hemodynamic measurements are shown for the overall analytic cohort and stratified by adjudicated G2PH and non G2PH. Continuous variables are reported as median [interquartile range] and categorical variables as n (%). P values reflect unadjusted comparisons between G2PH and non G2PH participants using Wilcoxon rank-sum tests for continuous variables and χ² or Fisher exact tests for categorical variables, as appropriate. **Abbreviations:** BMI, body mass index; CAD, coronary artery disease; DPG, diastolic pressure gradient; FC, fluid challenge; HFpEF, heart failure with preserved ejection fraction; HFrEF, heart failure with reduced ejection fraction; iCPET, invasive cardiopulmonary exercise testing; iNO, inhaled nitric oxide; LAVI, left atrial volume index; LVEF, left ventricular ejection fraction; mPAP, mean pulmonary artery pressure; PCWP, pulmonary capillary wedge pressure; PH, pulmonary hypertension; PVR, pulmonary vascular resistance; RAP, right atrial pressure; RVFAC, right ventricular fractional area change; RV/LV, right ventricular–to–left ventricular ratio; TAPSE, tricuspid annular plane systolic excursion; TPG, transpulmonary gradient; VO₂, oxygen consumption. *, †, ‡, §, ||, #, **

### Discordant Phenotype–Hemodynamic Profiles Demonstrate Distinct Physiologic Signatures

Despite these expected global physiologic differences, substantial overlap persisted between adjudicated phenotypes across resting PCWP ranges (**Figure 3**). Concordant and discordant phenotype–hemodynamic groups demonstrated distinct but internally consistent physiologic profiles across testing stages.

**Figure 3.**
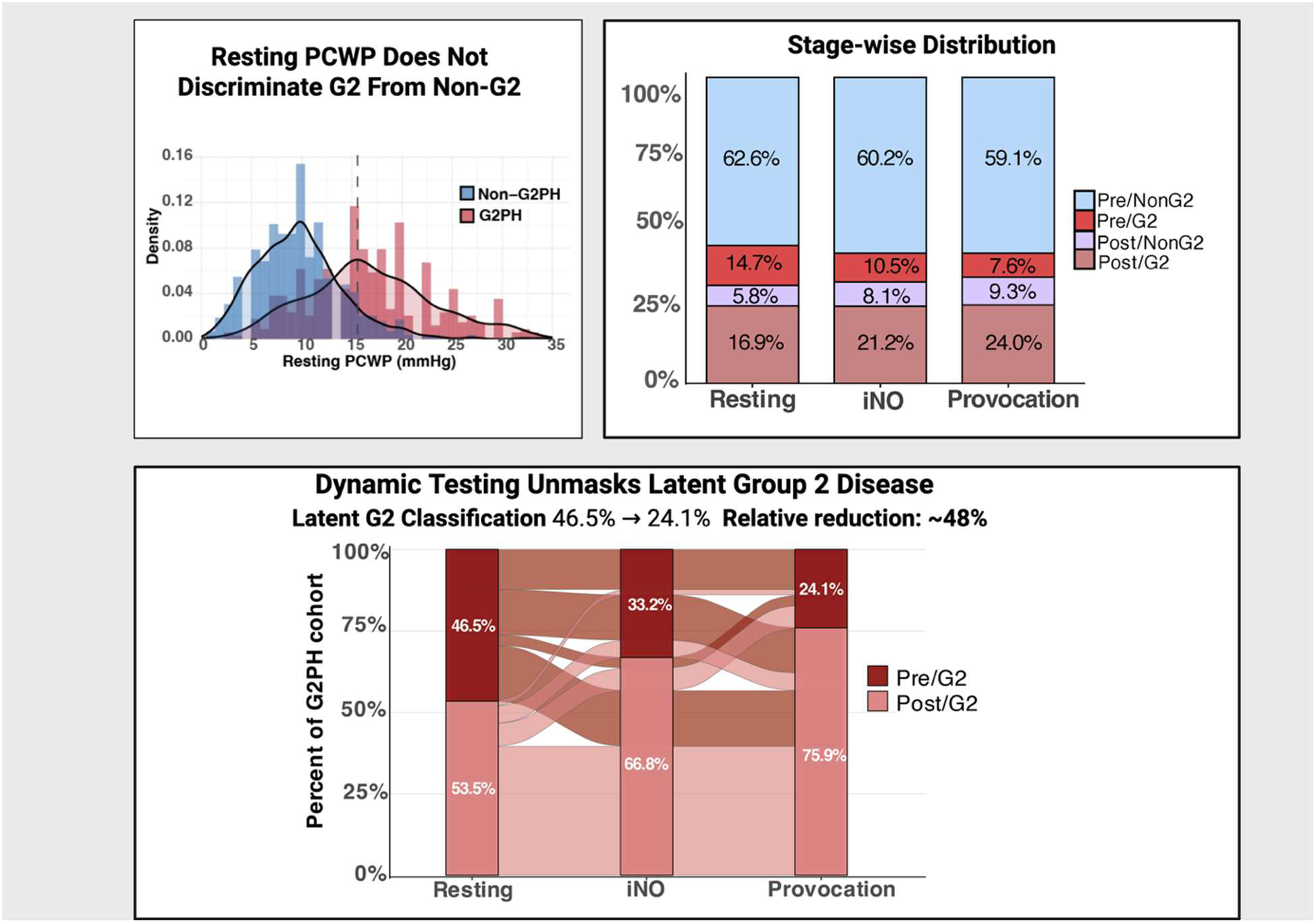
Stage-wise reclassification demonstrates reduction in latent Group 2 PH. **Upper left:** Distribution of resting pulmonary capillary wedge pressure (PCWP) in adjudicated Group 2 and Non–Group 2 participants demonstrate substantial overlap across the intermediate range, illustrating why single-threshold classification results in discordant phenotypes. **Upper right:** Distribution of concordance categories at rest, after inhaled nitric oxide (iNO), and after provocation (fluid challenge or invasive cardiopulmonary exercise testing). The proportion of latent Group 2 (Pre/G2) decreases across stages, while true positive (Post/G2) classification increases. **Lower panel:** Alluvial plot illustrating reclassification of Group 2 subjects across staged testing. At rest, 46.5% of Group 2 subjects were misclassified as latent Group 2 (Pre/G2). Following iNO, this decreased to 33.2%, and after provocation to 24.1%, with corresponding increases in true positive classification. **Abbreviations:** G2PH, Group 2 pulmonary hypertension; iNO, inhaled nitric oxide; Prov, provocation (fluid challenge or exercise); WSPH, World Symposium on Pulmonary Hypertension.

Relative to Pre/NonG2 participants, Pre/G2 participants exhibited higher baseline PCWP, greater augmentation in PCWP during iNO and provocation, and progressive increases in RAP with dynamic loading, consistent with impaired left-sided reserve physiology (**Supplemental Tables S3-7).** In contrast, Post/NonG2 participants retained higher transpulmonary gradients, pulmonary vascular loading, and RV remodeling despite elevated resting wedge pressure, supporting a pulmonary vascular/interdependence phenotype rather than intrinsic left-heart disease alone.

Stage-wise distributions of concordance groups shifted progressively across physiologic testing stages (**Figure 3**). The proportion of Pre/G2 participants decreased from 14.7% at rest to 10.5% following iNO and 7.6% following provocation, whereas Post/G2 concordance increased from 16.9% to 21.2% and 24.0%, respectively. Among adjudicated Group 2 participants, the proportion classified as Pre/G2 decreased from 46.5% at rest to 33.2% following iNO and 24.1% following provocation, while Post/G2 concordance increased from 53.5% to 66.8% and 75.9%, respectively.

For staged analyses, exercise and fluid challenge were grouped as a unified provocation stage to evaluate the incremental contribution of dynamic reserve physiology beyond resting and iNO assessment. This approach was based on their shared physiologic objective of increasing LV filling requirements and on directionally similar effects on phenotype–hemodynamic concordance (**Supplemental Tables S5-8**).

### Ventricular Interdependence Contributes to Discordant Post-Capillary Physiology

Discordant post-capillary physiology (Post/NonG2) demonstrated structural and hemodynamic features consistent with ventricular interdependence (**Figure 4**). Transmural pressure differed significantly across concordance groups overall (p<0.001), but Post/NonG2 participants demonstrated transmural pressures like Post/G2 participants (p=0.223), despite distinct adjudicated phenotypes. In contrast, RV/LV ratio was higher in Post/NonG2 participants compared with Post/G2 participants (p<0.001).

**Figure 4.**
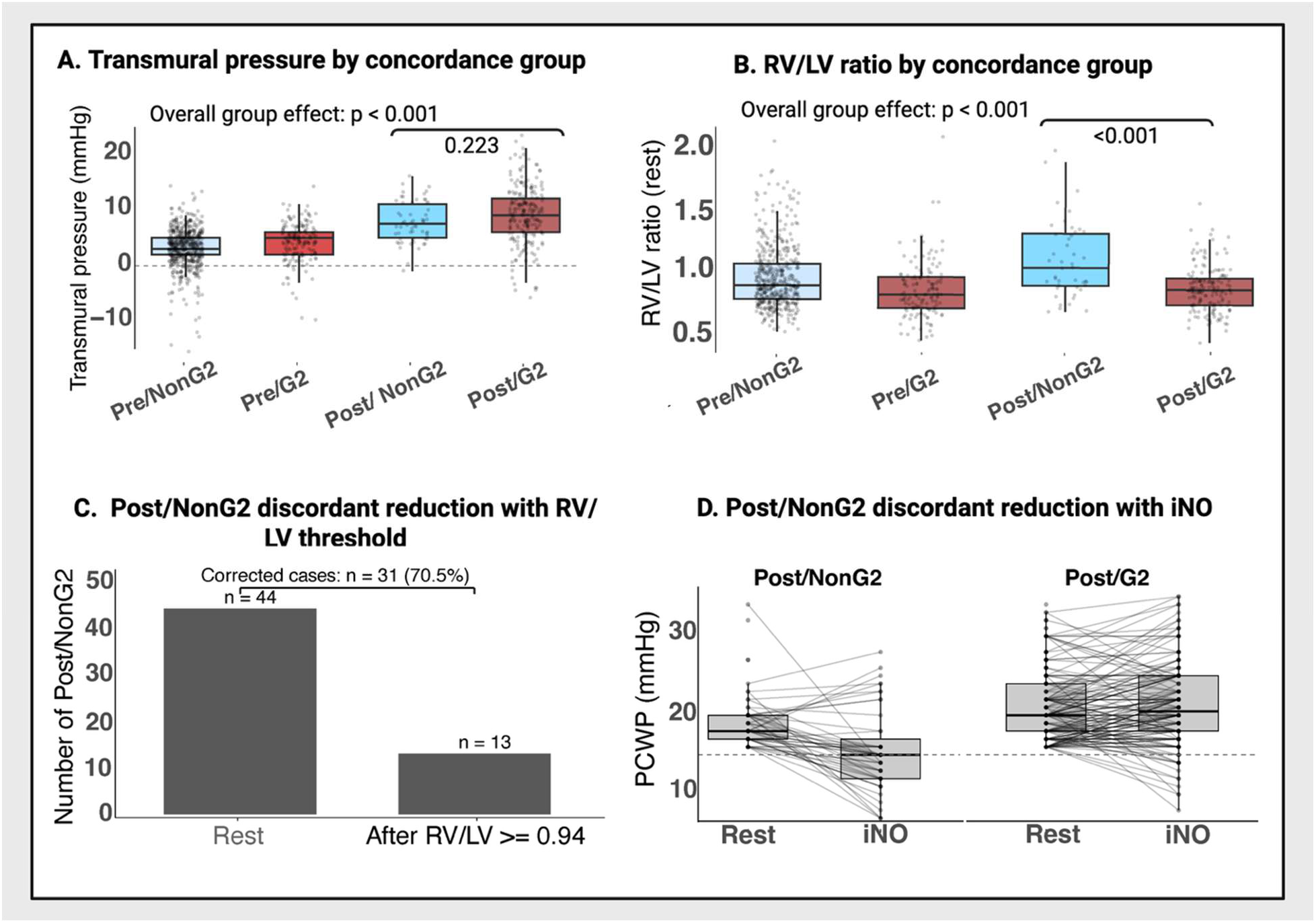
Pericardial constraint, ventricular interdependence, and false-positive post-capillary classification. **Panel A:** Resting transmural pressure by concordance group. Although overall differences were observed (p < 0.001), transmural pressure did not distinguish discordant post-capillary physiology (Post/NonG2) from true Group 2 PH (Post/G2). **Panel B:** Echocardiographic right ventricular–to–left ventricular (RV/LV) diastolic diameter ratio by concordance group. RV/LV differed across groups (p < 0.001) and was higher in discordant Post/NonG2 compared with true Group 2 (Post/G2) (p < 0.001). **Panel C:** Reduction in discordant classification using RV/LV ≥0.94. Among discordant cases at rest (Post/NonG2, n=44), 31 (70.5%) were reclassified as non–Group 2, indicating improved specificity. **Panel D:** Change in PCWP with inhaled nitric oxide (iNO). Discordant Post/NonG2 cases show a reduction in PCWP, whereas true Group 2 cases demonstrate minimal change or persistent elevation. **Abbreviations:** iNO, inhaled nitric oxide; PCWP, pulmonary capillary wedge pressure; RV/LV, right ventricular–to–left ventricular ratio.

Application of an RV/LV ratio threshold ≥0.94 reduced discordant Post/NonG2 profiles by 70.5%, from 44 to 13 participants (**Figure 4**). During iNO, Post/NonG2 participants demonstrated reductions in PCWP, whereas Post/G2 participants maintained persistently elevated wedge pressures (**Figure 4 and Supplemental Figure S3, Table S9**).

### Intermediate Resting PCWP Ranges Demonstrate Bidirectional Phenotype–Hemodynamic Discordance

Resting PCWP demonstrated strong overall concordance with adjudicated phenotype assignment (AUC 0.86) but substantial bidirectional phenotype–hemodynamic overlap persisted across intermediate resting PCWP ranges (**Supplemental Table S10**; **Figure 3, upper left**). No discrete inflection points separated phenotypes, and spline analyses demonstrated continuous transition across the intermediate range without a clear biologic threshold (**Supplemental Figure S4**).

Resting PCWP thresholds associated with high phenotype–hemodynamic concordance clustered at physiologic extremes, whereas intermediate PCWP values represented overlapping probabilities of discordance rather than discrete biologic states (**Figure 5, left panel).** For example, resting PCWP values within the recommended intermediate range of 12–18 mmHg^1,8^ demonstrated persistent bidirectional phenotype–hemodynamic discordance. At a resting PCWP of 12 mmHg, 25% of participants classified as pre-capillary were Group 2 PH, whereas at PCWP values at 18 mmHg, 35% of participants classified as post-capillary remained discordant Post/NonG2. Phenotype–hemodynamic concordance did not approach 90% until resting PCWP values were <9 mmHg or >24 mmHg.

**Figure 5.**
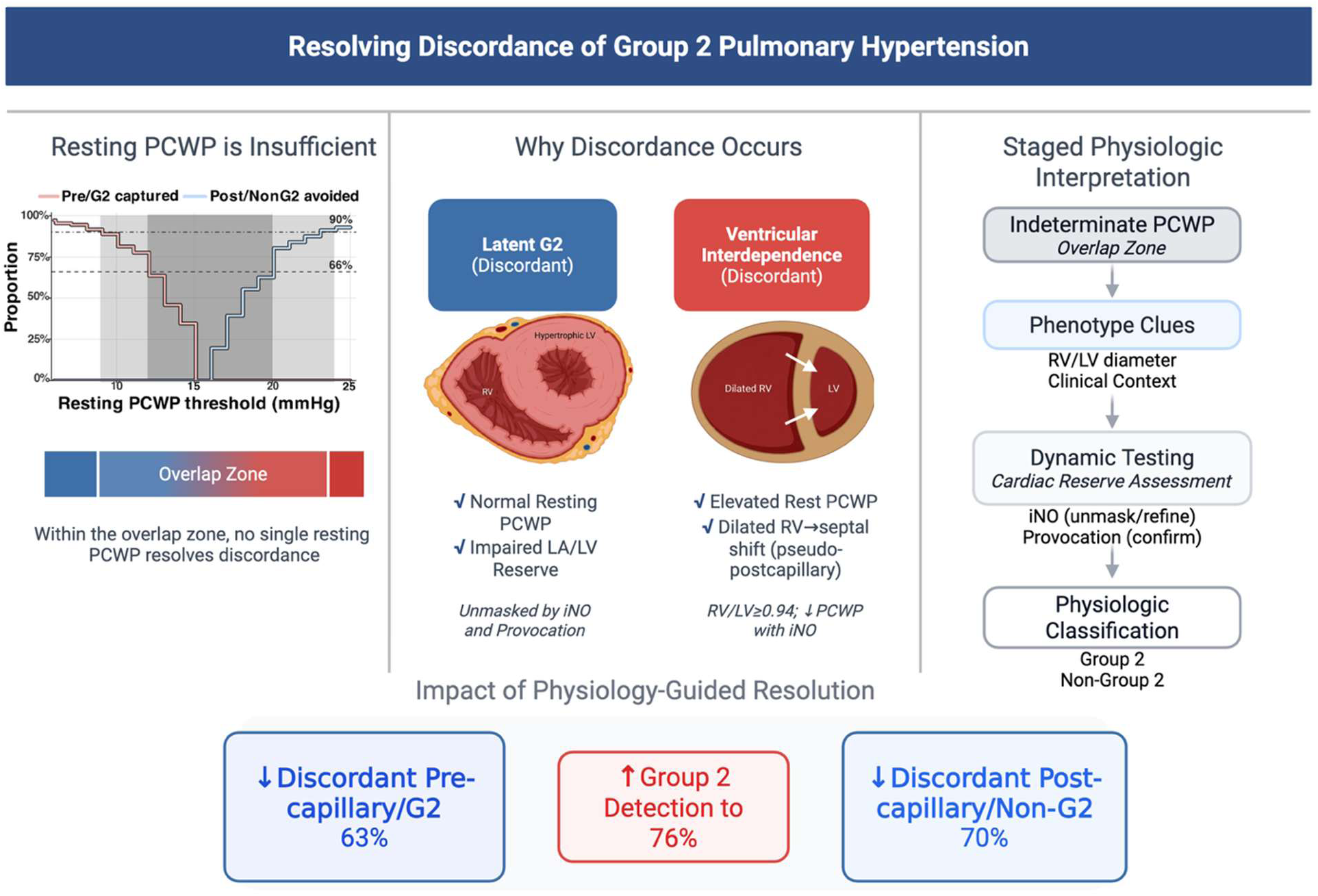
Physiology-guided resolution of phenotype–hemodynamic discordance in Group 2 pulmonary hypertension. **Left panel:** Resting pulmonary capillary wedge pressure (PCWP) demonstrates broad overlap between adjudicated Group 2 and non–Group 2 PHs, particularly within intermediate PCWP ranges, indicating that no single resting threshold fully resolves phenotype–hemodynamic discordance. Two reference thresholds for concordance certainty are shown schematically. Dark grey shading represents an intermediate-certainty zone (∼66–75% concordance), whereas light grey shading represents higher-certainty thresholds (∼90% concordance), corresponding approximately to resting PCWP values <9 mmHg and >24 mmHg. **Middle panel:** Two bidirectional discordance mechanisms are illustrated. Latent Group 2 PH (discordant Pre/G2) reflects impaired left atrial/left ventricular reserve despite normal or borderline resting PCWP and may be unmasked during dynamic testing. Discordant Post/NonG2 physiology reflects elevated resting PCWP despite absence of intrinsic left-heart disease, potentially related to ventricular interdependence, right ventricular dilation, septal shift, and pericardial restraint. **Right panel:** Proposed staged physiology-guided framework for resolving indeterminate resting PCWP profiles. Clinical phenotype clues inhaled nitric oxide (iNO), and provocative testing (exercise or fluid challenge) provide incremental physiologic context to refine classification relative to adjudicated phenotype assignment. A more detailed staged diagnostic framework is provided in Supplemental Figure S7. Overall, staged testing reduced discordant Pre/G2 profiles by 63%, reduced discordant Post/NonG2 profiles by 70%, and increased identification of adjudicated Group 2 PH to 83.7%.

### Sensitivity Analysis Using End-Expiratory PCWP

End-expiratory PCWP was systematically higher than respiratory averaged (mean) PCWP (**Supplemental Figure S5-6**). This resulted in up classification from precapillary to postcapillary in 138 (14.6%) of subjects. Sensitivity analyses using end-expiratory PCWP demonstrated higher sensitivity but lower specificity at rest, whereas respiratory-averaged PCWP showed the opposite pattern (**Supplemental Table S11**). End-expiratory measurements reduced Pre/G2 discordance while increasing Post/NonG2 discordance, indicating that respiratory gating modified sensitivity-specificity tradeoffs but did not eliminate intermediate phenotype–hemodynamic overlap. Despite these differences in resting operating characteristics, iNO and provocative testing produced similar directional changes across stages for both waveform methodologies, suggesting that staged physiologic responses were preserved independent of waveform selection (**Supplemental Table S11**).

### Physiology-Guided Staged Evaluation

Among subjects without resting PH, only 3 of 63 (5%) participants with adjudicated left heart disease had resting PCWP >15 mmHg. Provocative testing identified an additional 13 cases according to stage-specific PCWP thresholds, increasing detection to 16 of 63 (25%). Although this represented a statistically significant improvement over resting assessment alone (LR p-value <0.001), overall discrimination remained modest (AUC 0.59; sensitivity 25.4%), suggesting limited yield of provocative testing in the absence of PH.

In contrast, among subjects with resting PH, staged physiologic testing produced progressive improvements in phenotype–hemodynamic concordance (**Figure 5 and Supplemental Figure S7**). Resting PCWP values at physiologic extremes demonstrated high concordance with adjudicated G2PH phenotype, whereas intermediate PCWP ranges were enriched for both latent Group 2 disease and discordant post-capillary physiology.

Nested staged analyses demonstrated progressive increases in AUC, sensitivity, likelihood ratios, and overall classification performance across testing stages (**Supplemental Table S10**). Addition of RV/LV ratio and iNO response improved discrimination beyond resting PCWP alone, while incorporation of provocative testing further reduced discordant Pre/G2 classification and increased cumulative identification of adjudicated Group 2 PH (**Supplemental Table S12**). Among exercise-provoked participants, incorporation of PCWP/CO slope appeared to improve physiologic specificity for Group 2 PH by reducing discordant Post/NonG2 classification during exertion (**Supplemental Table S12**). Exploratory unconstrained CART analyses independently identified provocative testing responses as the dominant discriminator of Group 2 PH (**Supplemental Figure S8**).

Modality-specific staged analyses demonstrated directionally similar improvement in phenotype–hemodynamic concordance across both exercise and fluid challenge paradigms (**Supplemental Tables S13-14**). Exercise testing demonstrated greater sensitivity and greater reduction in discordant Pre/G2 classification, whereas fluid challenge demonstrated higher specificity. Both modalities incrementally increased adjudicated Group 2 PH identification beyond resting and iNO assessment.

Importantly, residual discordance persisted despite staged testing, indicating that no single hemodynamic threshold or perturbational strategy fully resolved overlap between clinically integrated phenotype and invasive hemodynamic profile (**Supplemental Table S15**). Compared with the broader Group 2 cohort, these individuals had lower body mass index, less hypertension and HFpEF, and a higher prevalence of valvular heart disease.

## Discussion

In this deeply phenotyped, protocolized PVDOMICS cohort, resting pulmonary capillary wedge pressure (PCWP) demonstrated substantial overall discriminatory value for adjudicated Group 2 PH, yet clinically important discordance persisted across intermediate pressure ranges. These findings support the concept that Group 2 pulmonary hypertension (PH) is fundamentally a clinically integrated phenotype rather than a purely hemodynamic state. Accordingly, pre-versus post-capillary classification should be interpreted within the broader physiologic and clinical context to refine diagnostic certainty and reduce misclassification.

A major implication of the present findings is that dynamic testing may be applied in a more targeted and physiologically informed manner rather than uniformly across all patients. In our data, intermediate resting PCWP values represented continuous probabilities of discordance rather than discrete diagnostic boundaries, permitting uncertainty thresholds to be quantified across the pressure spectrum. For example, resting PCWP values <9 mmHg and >24 mmHg demonstrated approximately 90% concordance with adjudicated phenotype assignment, whereas intermediate ranges carried substantially greater bidirectional uncertainty. Within these overlap zones, staged testing provided quantifiable incremental diagnostic yield, with iNO offering a practical low-burden intermediate step that improved classification before escalation to provocative testing. Because iNO is readily available during most diagnostic catheterizations, this nested approach may improve feasibility across centers where routine exercise or fluid challenge testing is less practical. However, exploratory CART analyses independently identified provocation as the single strongest discriminator of adjudicated Group 2 PH, supporting its use when a single dynamic test strategy is pursued. A more detailed staged physiology-guided framework is provided in **Supplemental Figure S7**.

These findings extend prior studies evaluating provocative testing in suspected HFpEF and pulmonary vascular disease. Previous investigations demonstrated that exercise and fluid loading may uncover occult post-capillary physiology, but most relied on PCWP-based criteria as both the diagnostic test and reference standard.^18,24,25^ The present analysis differs in several important respects. First, classification was benchmarked against independent multidisciplinary adjudication integrating clinical, imaging, physiologic, and hemodynamic information rather than wedge pressure alone. Second, participants underwent standardized sequential testing across multiple physiologic stages, permitting direct quantification of incremental diagnostic yield and false-positive burden. Third, the analysis characterized distinct discordant phenotypes rather than treating misclassification as a uniform entity.

Dynamic testing was particularly informative in participants with latent Group 2 PH. These individuals frequently demonstrated borderline resting PCWP despite impaired left atrial and left ventricular reserve that became apparent during physiologic stress. Exercise and fluid challenge produced directionally similar improvements in concordance, although exercise demonstrated greater sensitivity whereas fluid challenge demonstrated greater specificity. These findings support the physiologic relevance of dynamic reserve assessment independent of provocative modality and align with prior observations that impaired diastolic reserve may remain occult under resting conditions.^3,7,18,24,25^ Among exercise-provoked participants, incorporation of PCWP/CO slope appeared to further improve physiologic specificity by reducing discordance among patients exceeding position-appropriate exercise PCWP thresholds, suggesting that pressure–flow relationships may help distinguish flow-mediated wedge augmentation from disproportionate left-sided pressure responses during exertion. This physiologic application of PCWP/CO slope has not previously been systematically evaluated in the context of resolving phenotype–hemodynamic discordance and may help contextualize how exercise slope metrics can be integrated into clinical practice.

Inhaled nitric oxide (iNO) demonstrated a distinct physiologic profile that differed mechanistically from traditional provocative testing. Rather than directly increasing preload, iNO reduced right ventricular afterload and increased pulmonary blood flow.^26^ In latent Group 2 patients, this increase in flow unmasked impaired left-sided reserve and increased PCWP. In contrast, among discordant Post/NonG2 participants, iNO frequently reduced PCWP, suggesting that apparent post-capillary physiology was load-dependent and reversible rather than reflective of intrinsic left-heart disease. These bidirectional effects support the concept that iNO functions not simply as a pulmonary vasodilator test, but as a low-burden physiologic probe capable of both unmasking latent left-heart dysfunction and potentially resolving load-dependent false-positive classification.

The present findings also provide physiologic context to the ongoing controversy regarding end-expiratory versus respiratory-averaged PCWP measurement. Consistent with prior observations, end-expiratory PCWP increased sensitivity for adjudicated Group 2 PH across testing stages, but at the cost of reduced specificity and increased discordant post-capillary classification.^27^ Importantly, this tradeoff likely reflects the bidirectional nature of phenotype–hemodynamic discordance demonstrated in the current study. While higher PCWP measurements may improve detection of latent left-heart disease, elevated wedge pressure may also reflect ventricular interdependence and load-dependent filling abnormalities in advanced precapillary disease. These findings suggest that the optimal integration of respiratory methodology into staged diagnostic frameworks warrants further prospective study.

The physiologic basis of discordant Post/NonG2 classification appears linked to ventricular interdependence and pericardial constraint. These participants demonstrated marked RV-predominant geometry, elevated RV/LV ratio, and some showed reversible wedge elevation during iNO administration. In advanced precapillary disease, RV dilation and septal shift may distort LV geometry, impair LV filling, and elevate PCWP despite absence of intrinsic left ventricular pathology.^8,28,29^ Prior conductance catheter studies similarly demonstrated load-reversible LV stiffening in precapillary PH, contrasting with the intrinsic, load-insensitive stiffness characteristic of HFpEF.^30^ Consistent with this mechanism, false-positive post-capillary profiles clustered within intermediate PCWP ranges and improved substantially after incorporation of RV/LV geometry and dynamic testing. Together, these findings emphasize that wedge pressure should be interpreted within the context of biventricular structure and dynamic load responsiveness rather than as an isolated static measurement.

### Limitations

First, adjudicated Group 2 PH, although more clinically integrated than resting hemodynamics alone, remains an imperfect reference standard and may be susceptible to incorporation bias because PCWP contributed in part to adjudication. However, adjudication incorporated multidimensional clinical, structural, and physiologic data rather than wedge pressure alone, and similar directional findings were observed across concordance, staged, and high-certainty analyses.

Second, the staged framework was derived and evaluated within the same cohort and therefore requires external validation before broader clinical implementation. Although PVDOMICS provides an unusually standardized and deeply phenotyped dataset, replication in independent cohorts may be challenging.

Finally, fluid challenge and exercise testing were not uniformly performed in identical participants and were combined analytically as provocation because stage-wise physiologic and concordance trajectories were directionally similar. Accordingly, direct head-to-head comparisons between exercise and fluid challenge should be interpreted cautiously.

## Conclusions

Group 2 pulmonary hypertension is a dynamic, load-dependent phenotype that cannot be reliably characterized by resting hemodynamics alone. Intermediate PCWP values represent continuous probabilities of bidirectional phenotype–hemodynamic discordance rather than discrete diagnostic boundaries. Dynamic testing improves physiologic classification by unmasking latent left-heart disease while resolving load-dependent false-positive post-capillary physiology related to ventricular interdependence. Inhaled nitric oxide provides a unique low-burden physiologic probe that complements provocative testing through mechanistically distinct effects on pulmonary flow and ventricular loading. Collectively, these findings support a staged, physiology-guided framework for refining pulmonary hypertension classification in contemporary clinical practice.

## Data Availability

Data is available upon reasonable request provided that it complies with the PVDOMICS steering committee rules and regulations.

## Non-standard abbreviations

ANOVA: analysis of variance
AUC: area under the receiver operating characteristic curve
BMI: body mass index
CAD: coronary artery disease
CART: classification and regression tree
CI: confidence interval
CPET: cardiopulmonary exercise testing
DPG: diastolic pressure gradient
FC: fluid challenge
FN: false negative
FP: false positive
G2: Group 2 disease
G2PH: Group 2 pulmonary hypertension
HFpEF: heart failure with preserved ejection fraction
HFrEF: heart failure with reduced ejection fraction
iCPET: invasive cardiopulmonary exercise testing
iNO: inhaled nitric oxide
LA: left atrium
LAVI: left atrial volume index
LHD: left heart disease
LV: left ventricle
LVEF: left ventricular ejection fraction
mPAP: mean pulmonary artery pressure
PA: pulmonary artery
PCWP: pulmonary capillary wedge pressure
PH: pulmonary hypertension
PVR: pulmonary vascular resistance
RAP: right atrial pressure
ROC: receiver operating characteristic
RV: right ventricle
RVFAC: right ventricular fractional area change
RV/LV: echocardiographic right ventricular–to–left ventricular diastolic ratio
SpO₂: peripheral oxygen saturation
TMP: transmural pressure
TN: true negative
TP: true positive
TPG: transpulmonary gradient
VO₂: peak oxygen consumption
WSPH: World Symposium on Pulmonary Hypertension

## References

1. Kovacs G, Bartolome S, Denton CP, Gatzoulis MA, Gu S, Khanna D, Badesch D, Montani D. Definition, classification and diagnosis of pulmonary hypertension. Eur Respir J. 2024. doi: 10.1183/13993003.01324-2024

2. Weatherald J, Hemnes AR, Maron BA, Mielniczuk LM, Gerges C, Price LC, Hoeper MM, Humbert M. Phenotypes in pulmonary hypertension. Eur Respir J. 2024;64. doi: 10.1183/13993003.01633-2023

3. Maron BA, Bortman G, De Marco T, Huston JH, Lang IM, Rosenkranz SH, Vachiery JL, Tedford RJ. Pulmonary hypertension associated with left heart disease. Eur Respir J. 2024. doi: 10.1183/13993003.01344-2024

4. Hoeper MM, Oerke B, Wissmüller M, Leuchte H, Opitz C, Halank M, Seyfarth HJ, Baldus S, Bauersachs J, Böhm M, et al. Tadalafil for Treatment of Combined Postcapillary and Precapillary Pulmonary Hypertension in Patients With Heart Failure and Preserved Ejection Fraction: A Randomized Controlled Phase 3 Study. Circulation. 2024;150:600–610. doi: 10.1161/circulationaha.124.069340

5. Bermejo J, Yotti R, García-Orta R, Sánchez-Fernández PL, Castaño M, Segovia-Cubero J, Escribano-Subías P, San Román JA, Borrás X, Alonso-Gómez A, et al. Sildenafil for improving outcomes in patients with corrected valvular heart disease and persistent pulmonary hypertension: a multicenter, double-blind, randomized clinical trial. Eur Heart J. 2018;39:1255–1264. doi: 10.1093/eurheartj/ehx700

6. Vachiery JL, Delcroix M, Al-Hiti H, Efficace M, Hutyra M, Lack G, Papadakis K, Rubin LJ. Macitentan in pulmonary hypertension due to left ventricular dysfunction. Eur Respir J. 2018;51. doi: 10.1183/13993003.01886-2017

7. Maron BA, Kovacs G, Vaidya A, Bhatt DL, Nishimura RA, Mak S, Guazzi M, Tedford RJ. Cardiopulmonary Hemodynamics in Pulmonary Hypertension and Heart Failure: JACC Review Topic of the Week. J Am Coll Cardiol. 2020;76:2671–2681. doi: 10.1016/j.jacc.2020.10.007

8. Rayner SG, Tedford RJ, Leary PJ, Mak S, Houston BA. “This Patient Needs a Doctor, Not a Guideline!” The Zone of Uncertainty in Pulmonary Arterial Wedge Pressure Measurement. Am J Respir Crit Care Med. 2024;210:712–714. doi: 10.1164/rccm.202402-0359VP

9. Hoeper MM. Navigating Uncertainty in PAH Classification: Wedge Pressure Versus Disease Phenotypes. Circulation. 2025;152:4–6. doi: 10.1161/CIRCULATIONAHA.125.073885

10. Rosenkranz S, Gibbs JS, Wachter R, De Marco T, Vonk-Noordegraaf A, Vachiery JL. Left ventricular heart failure and pulmonary hypertension. Eur Heart J. 2016;37:942–954. doi: 10.1093/eurheartj/ehv512

11. Borlaug BA, Nishimura RA, Sorajja P, Lam CS, Redfield MM. Exercise hemodynamics enhance diagnosis of early heart failure with preserved ejection fraction. Circ Heart Fail. 2010;3:588–595. doi: 10.1161/CIRCHEARTFAILURE.109.930701

12. Hemnes AR, Beck GJ, Newman JH, Abidov A, Aldred MA, Barnard J, Berman Rosenzweig E, Borlaug BA, Chung WK, Comhair SAA, et al. PVDOMICS: A Multi-Center Study to Improve Understanding of Pulmonary Vascular Disease Through Phenomics. Circulation Research. 2017;121:1136–1139. doi: 10.1161/CIRCRESAHA.117.311737

13. Hemnes AR, Leopold JA, Radeva MK, Beck GJ, Abidov A, Aldred MA, Barnard J, Rosenzweig EB, Borlaug BA, Chung WK, et al. Clinical Characteristics and Transplant-Free Survival Across the Spectrum of Pulmonary Vascular Disease. J Am Coll Cardiol. 2022;80:697–718. doi: 10.1016/j.jacc.2022.05.038

14. Kovacs G, Herve P, Barbera JA, Chaouat A, Chemla D, Condliffe R, Garcia G, Grunig E, Howard L, Humbert M, et al. An official European Respiratory Society statement: pulmonary haemodynamics during exercise. Eur Respir J. 2017;50:1–18. doi: 10.1183/13993003.00578-2017

15. Tedford RJ, Houston BA, Jackson GR, Viray MC, Bonno EL. Modern Right Heart Catheterization: Beyond Simple Hemodynamics. Advances in Pulmonary Hypertension. 2020;19:6–15. doi: 10.21693/1933-088x-19.1.6

16. Borlaug BA, Reddy YNV. The Role of the Pericardium in Heart Failure: Implications for Pathophysiology and Treatment. JACC Heart Fail. 2019;7:574–585. doi: 10.1016/j.jchf.2019.03.021

17. Borlaug BA, Sharma K, Shah SJ, Ho JE. Heart Failure With Preserved Ejection Fraction: JACC Scientific Statement. J Am Coll Cardiol. 2023;81:1810–1834. doi: 10.1016/j.jacc.2023.01.049

18. Montane B, Tonelli AR, Arunachalam A, Bhattacharyya A, Li M, Wang X, Chaisson NF. Hemodynamic Responses to Provocative Maneuvers during Right Heart Catheterization. Ann Am Thorac Soc. 2022;19:1977–1985. doi: 10.1513/AnnalsATS.202201-077OC

19. Oliveira RK, Agarwal M, Tracy JA, Karin AL, Opotowsky AR, Waxman AB, Systrom DM. Age-related upper limits of normal for maximum upright exercise pulmonary haemodynamics. Eur Respir J. 2016;47:1179–1188. doi: 10.1183/13993003.01307-2015

20. Esfandiari S, Wolsk E, Granton D, Azevedo L, Valle FH, Gustafsson F, Mak S. Pulmonary Arterial Wedge Pressure at Rest and During Exercise in Healthy Adults: A Systematic Review and Meta-analysis. J Card Fail. 2019;25:114–122. doi: 10.1016/j.cardfail.2018.10.009

21. Eisman AS, Shah RV, Dhakal BP, Pappagianopoulos PP, Wooster L, Bailey C, Cunningham TF, Hardin KM, Baggish AL, Ho JE, et al. Pulmonary Capillary Wedge Pressure Patterns During Exercise Predict Exercise Capacity and Incident Heart Failure. Circ Heart Fail. 2018;11:e004750. doi: 10.1161/CIRCHEARTFAILURE.117.004750

22. Eisman AS, Shah RV, Dhakal BP, Pappagianopoulos PP, Wooster L, Bailey C, Cunningham TF, Hardin, Kathryn M., Baggish AL, Ho JE, et al. Pulmonary Capillary Wedge Pressure Patterns During Exercise Predict Exercise Capacity and Incident Heart Failure. Circulation: Heart Failure. 2018;11:e004750. doi: doi:10.1161/CIRCHEARTFAILURE.117.004750

23. DeLong ER, DeLong DM, Clarke-Pearson DL. Comparing the Areas under Two or More Correlated Receiver Operating Characteristic Curves: A Nonparametric Approach. Biometrics. 1988;44:837–845. doi: 10.2307/2531595

24. Guazzi M, Naeije R. Pulmonary Hypertension in Heart Failure: Pathophysiology, Pathobiology, and Emerging Clinical Perspectives. J Am Coll Cardiol. 2017;69:1718–1734. doi: 10.1016/j.jacc.2017.01.051

25. Humbert M, Kovacs G, Hoeper MM, Badagliacca R, Berger RMF, Brida M, Carlsen J, Coats AJS, Escribano-Subias P, Ferrari P, et al. 2022 ESC/ERS Guidelines for the diagnosis and treatment of pulmonary hypertension. Eur Heart J. 2022;43:3618–3731. doi: 10.1093/eurheartj/ehac237

26. Krishtopaytis E, Ampnti SA, Obeidat M, Ramahi N, Lane J, Toth D, Paul D, Tonelli AR. Can Inhaled Nitric Oxide Response Predict Tolerance to Therapies and Survival in Patients With Combined Precapillary and Postcapillary Pulmonary Hypertension? Am J Cardiol. 2023;207:363–369. doi: 10.1016/j.amjcard.2023.09.032

27. LeVarge BL, Pomerantsev E, Channick RN. Reliance on end-expiratory wedge pressure leads to misclassification of pulmonary hypertension. Eur Respir J. 2014;44:425–434. doi: 10.1183/09031936.00209313

28. Naeije R, Badagliacca R. The overloaded right heart and ventricular interdependence. Cardiovasc Res. 2017;113:1474–1485. doi: 10.1093/cvr/cvx160

29. Blanchard DG, Dittrich HC. Pericardial adaptation in severe chronic pulmonary hypertension. An intraoperative transesophageal echocardiographic study. Circulation. 1992;85:1414–1422. doi: 10.1161/01.cir.85.4.1414

30. Kasner M, Westermann D, Steendijk P, Drose S, Poller W, Schultheiss HP, Tschope C. Left ventricular dysfunction induced by nonsevere idiopathic pulmonary arterial hypertension: a pressure-volume relationship study. Am J Respir Crit Care Med. 2012;186:181–189. doi: 10.1164/rccm.201110-1860OC

